# Clinical Utility of Ultrasonography BI-RADS in the Evaluation of Breast Cancer in Patients with Palpable Breast Masses: A Diagnostic Test Accuracy Original Article

**DOI:** 10.1101/2023.11.08.23298253

**Authors:** Vismit Gami, Dev Desai, Sahil Shah, Devang Rana

**Affiliations:** Smt. NHLMMC, Ahmedabad, India; Department of Radiodiagnosis, Smt. NHLMMC, Ahmedabad, India; Department of Pharmacology, Smt. NHLMMC, Ahmedabad, India

**Keywords:** USG BI-RADS, Breast lump, Histopathology, Staging of Breast Carcinoma

## Abstract

**Introduction:** Diagnosing and staging breast cancer with an easy and widely useable method that can be employed worldwide in the poorest and wealthiest settings is important.

Mammography is a technique that might not be available in faraway clinics and it is technically challenging whereas USG can be available in most remote areas and small hospitals far from tertiary care hospitals. Even a trainee Radiology resident can use USG BIRADS and can be used diagnostically for that, it is important to define its, diagnostic accuracy with Sensitivity, Specificity, and other diagnostic parameters.

**Aims:** To determine the Diagnostic accuracy of USG BIRADS compared to the gold standard Histopathology report

**Methodology:** A Retrospective cohort study was conducted at a tertiary care hospital. a total of 84 female patients presenting to Surgical OPD with complaints of a breast lump or pain were enrolled from their records. Their Breast USG results were analyzed to identify their BIRADS stage correctly and then their corresponding Histopathology report was considered the gold standard to compare the USG results against. Excel, SPSS, and Revman were used to conduct analysis and create results.

**Results:** 36 of these 84 patients belonged to BIRADS 1, 2, and 5 where Sensitivity, Specificity, and PPV were calculated at 100%. No one was diagnosed with BIRADS III from USG reports. For USG BIRADS 4, in total 48 patients Sensitivity was 0.667, specificity was 0.883, and PPV was 0.364.

**Conclusion:** Patients whose USG shows Benign growth or can be diagnosed in BIRADS 1, 2, 3, and 5 can be counted as accurate and precise. When the USG diagnosis describes the patient to be in BIRADS 4, the sensitivity and PPV show poor results showing a very low probability of the patient being truly positive when the diagnosis gives a positive result.

## Introduction

Breast cancer is a complex disease that affects millions of female individuals worldwide with about 1 in every 8 females worldwide having a lifetime risk of developing breast cancer[1]. Significant progress has been made in the understanding, diagnosis, and treatment of breast cancer over the years. Breast cancer has ranked number one cancer among Indian females with age age-adjusted rate as high as 25.8 per 100,000 women and a mortality 12.7 per 100,000 women[2]. Triple assessment is an approach used in the diagnosis and management of breast cancer for any patients who present with a breast mass (lump) or any other complaint of breast suspicion of carcinoma. It involves three components that help us comprehensively evaluate breast abnormalities like lumps, mass, etc. The three components of triple assessment include clinical assessment, radiological imaging, and Histo-pathological analysis. The diagnosis should be made with a combination of clinical assessment, radiological imaging, and Histo-pathological analysis of the tissue sample. Triple assessment is considered positive when any one of the three modalities, physical examination of the breast, radiological imaging, and histo-pathological analysis is positive and is considered negative when all three modalities are negative[3].

Radiological imaging includes imaging techniques that give detailed visualization of the breast tissue. The most done imaging techniques are Mammography, Ultrasound Sonography, and Magnetic imaging resonance (MRI). Mammography is not routinely performed because of its ionizing radiation, and the decreased sensitivity of mammography in the frequently dense breasts of young women[4]. Mammography is a technique that might not be available in faraway clinics and it is technically challenging whereas USG can be available in most remote areas and small hospitals far from tertiary care hospitals. However, despite the widely accepted application of ultrasound in young women with focal signs or symptoms, there are sparse outcomes data to validate its use in this clinical setting and for that, it is important to define its, diagnostic accuracy with Sensitivity, Specificity, and other diagnostic parameters.

A standardized way of reporting Mammograms, USG, and MRIs is the BIRADS Score. Breast Imaging Recording and Data System (BIRADS) was developed by the American College of Radiology (ACR) to provide a uniform method for reporting mammography and breast imaging findings[5]. The BIRADS system uses a scale of categories ranging from 0 to 6[11], each indicating a specific level of suspicion or recommendation for further evaluation.

**Table 1:** Gross BI-RADS Classification.

| **Table 1:- Gross BI-RADS Classification** |  |
| --- | --- |
| 0 | Incomplete assessment or need for additional imaging evaluation |
| 1 | Negative or normal finding |
| 2 | Benign finding |
| 3 | Probably benign finding (follow-up recommended every 6 months) |
| 4 | Suspicious abnormality (biopsy recommended) |
| 5 | Highly suggestive of malignancy (biopsy strongly recommended) |
| 6 | Known biopsy-proven malignancy (treatment already underway). |

The BIRADS assessment is provided by a radiologist after reviewing the imaging results. It helps to standardize reporting across different healthcare facilities, referring physicians, and patients regarding the level of suspicion for breast abnormalities detected on imaging.

The purpose of this study was to determine the Diagnostic accuracy of USG BI-RADS compared to the gold standard Histopathology report [6]t for detecting malignancy when performed as the primary imaging test.

## Materials and Methodology

A Retrospective cohort study was conducted at a tertiary care hospital. A total of 84 female patients presenting to Surgical OPD with complaints of a breast lump or pain were enrolled from their records. Their Breast USG results were analyzed to correctly identify their BI-RADS stage. Then their corresponding Histopathology report was considered the gold standard to compare the USG results against. Excel, SPSS, and RevMan software were used to conduct analysis and create results.

### Inclusion Criteria

- Whose Breast USG data of breast lump is available?
- Whose Histopathology data for breast lesions is available.

### Exclusion Criteria

- Patients whose complete medical history is not available.
- Patients whose Breast USG or Histopathology of breast lesion has not been done by an expert Radiologist.

Diagnostic parameters like Sensitivity, Specificity, Odd’s Ratio, Youden Index, Negative Likelihood ratio, Positive likelihood ratio, PPV, NPV, and Area Under Curve for ROC were calculated along with frequency studies to calculate the results

## Imaging and Interpretation

All the ultrasounds were performed by radiologists that were specialized in breast imaging. With a 12Mhz linear transducer USG. The radiologist has described the lesion on the following characteristics: Breast side, location on the lesion, distance from the nipple, and imaging characteristics of sonographic imaging including lesion shape, margin, posterior acoustic features, echogenicity, and their correct BI-RADS scores were given after taking into consideration of all the following data. Corresponding Histo-pathological reports were also collected of the patients as gold standard, from the medical records of the patients.

## Results

A total of 84 patients were evaluated in our study and out of these Benign lesions accounted for 83% of all lesions recorded in our study and 17% were malignant lesions.

A predominance of fibroadenoma (68.6% of all benign cases), followed by inflammatory mastopathy like mastitis (11.4%).

Breast carcinoma was the chief Malignant lesion with 50% of all malignant lesions found, followed by Infiltrating Carcinoma (Ex-situ) with about 35.7%. Finally, 7.1% of the malignant lesions were Malignant DCIS (ductal carcinoma in situ) and IDC (invasive ductal carcinoma), respectively.

Out of 84 patients BI-RADS scoring was given by a radiologist based on Ultrasonography findings and the patients distribution is as follows:

20 Patients were identified as BI-RADS 1 followed by 14 patients in BI-RADS 2, 48 patients in BIRADS 4, and 2 patients in BIRADS 5. However, no patients were identified in BIRADS 3 in our study.

36 of these 84 patients belonged to BI-RADS 1, 2, and 5. The sensitivity, specificity, and Positive predictive value were calculated for BIRADS 1,2,5 and it was at 100%.

For USG BI-RADS 4, in total 48 patients Sensitivity was 66.7%, specificity was 88.3%, and PPV was 36.4%.

Other Statistical measures used in diagnostic testing to evaluate the performance and accuracy of USG BI-RADS 4. *Diagnostic Odd’s ratio* was at 10.28. *The Negative Likelihood ratio* was 0.25 and the *Positive Likelihood ratio* was 0.39.

The *Youden Index*[7] is calculated using the sensitivity (true positive rate) and specificity (true negative rate) of a test. It is defined as:

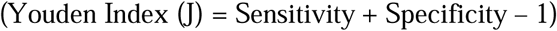

The index ranges from 0 to 1, with a higher value indicating a better test performance. A value of 0 indicates no discrimination ability of the test, while a value of 1 represents a perfect test. In our study, the Youden index was calculated and it was 0.5, which is in between.

To better understand the diagnostic value USG BI-RADS 4 *Receiver Operating characteristic (ROC)* curve was run and the results show Area under the curve for Receiver operative characteristic (ROC) is 0.7519.

AUC ranges from 0-1 where, the higher the AUC value, the better the performance of the modality at distinguishing between the positive and negative results. In our study, it was at 0.7519 which was the bare minimum for USG BIRADS 4.

**Fig. 1:**
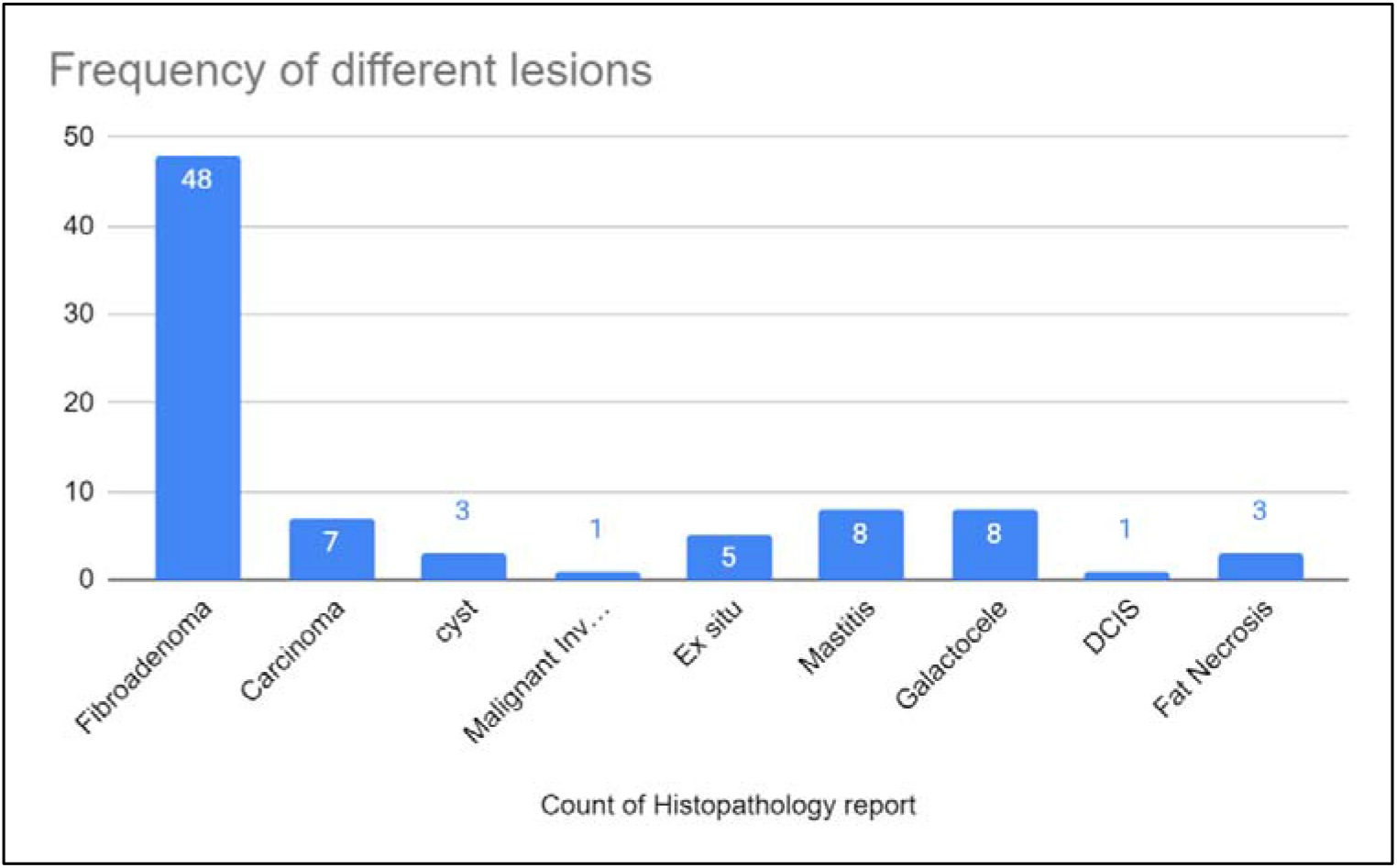
Frequency of different lesions.

**Fig. 2:**
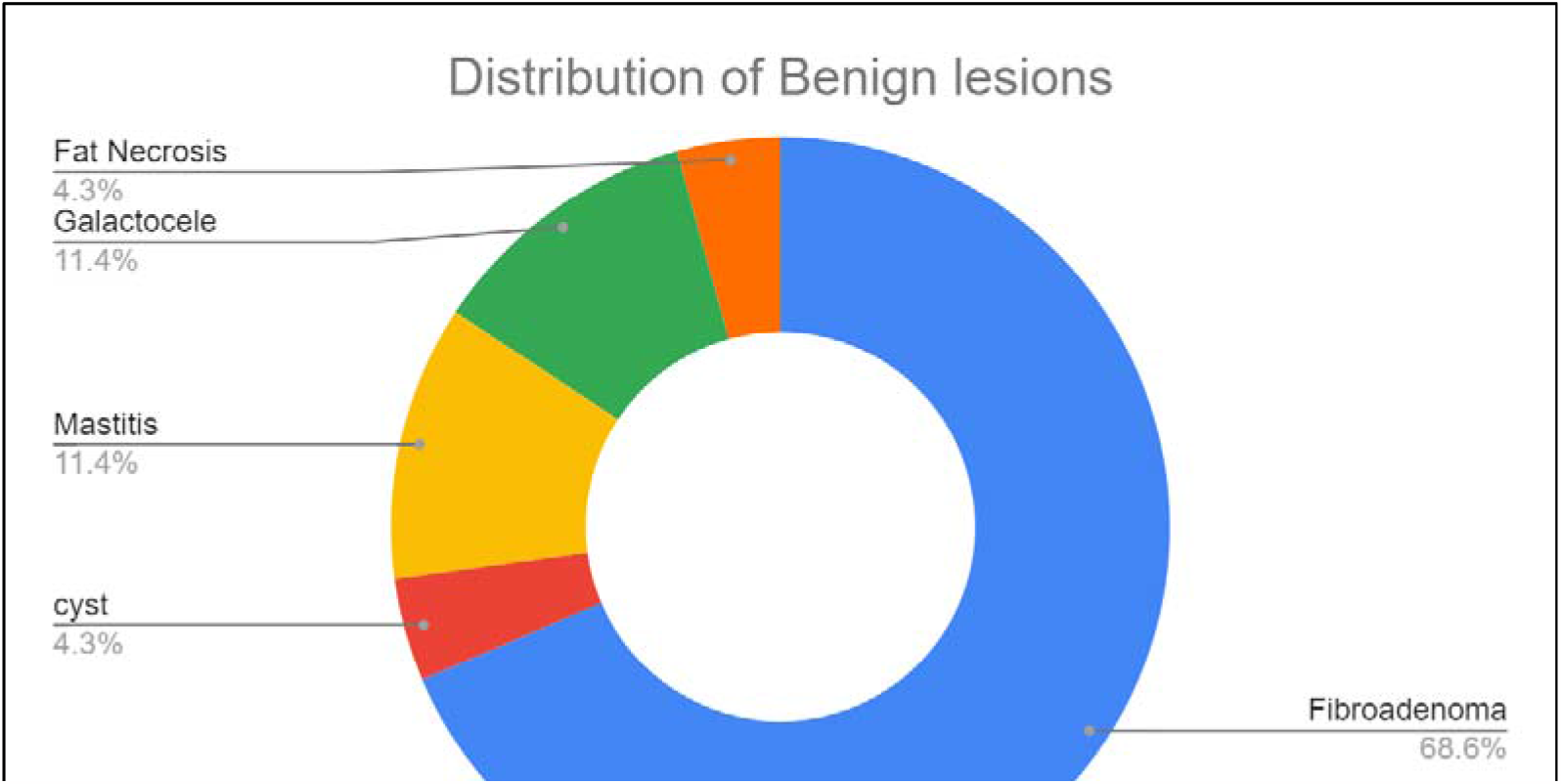
Distribution of Benign lesion.

**Fig. 3:**
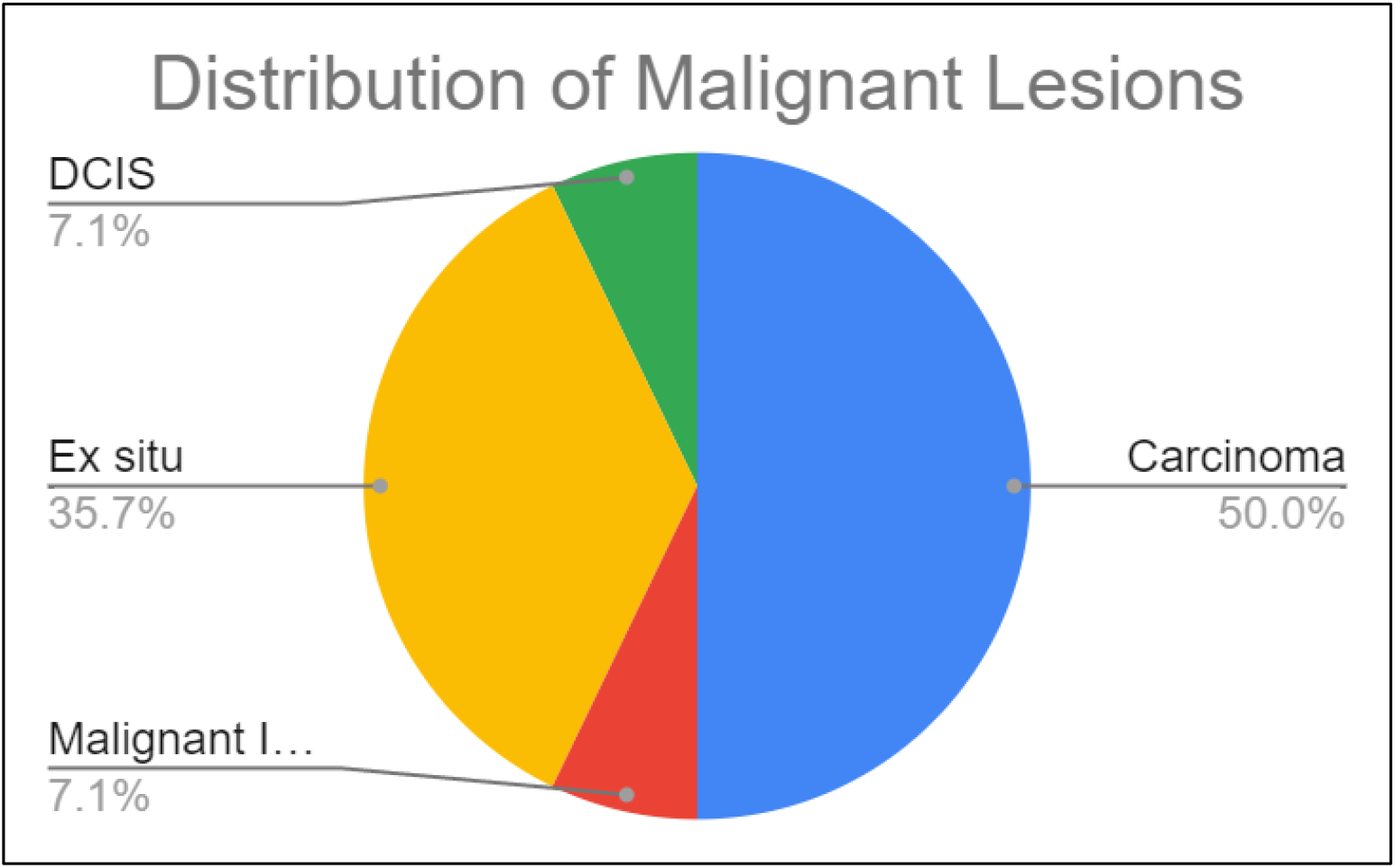
Distribution of Malignant lesion.

**Fig. 4.**
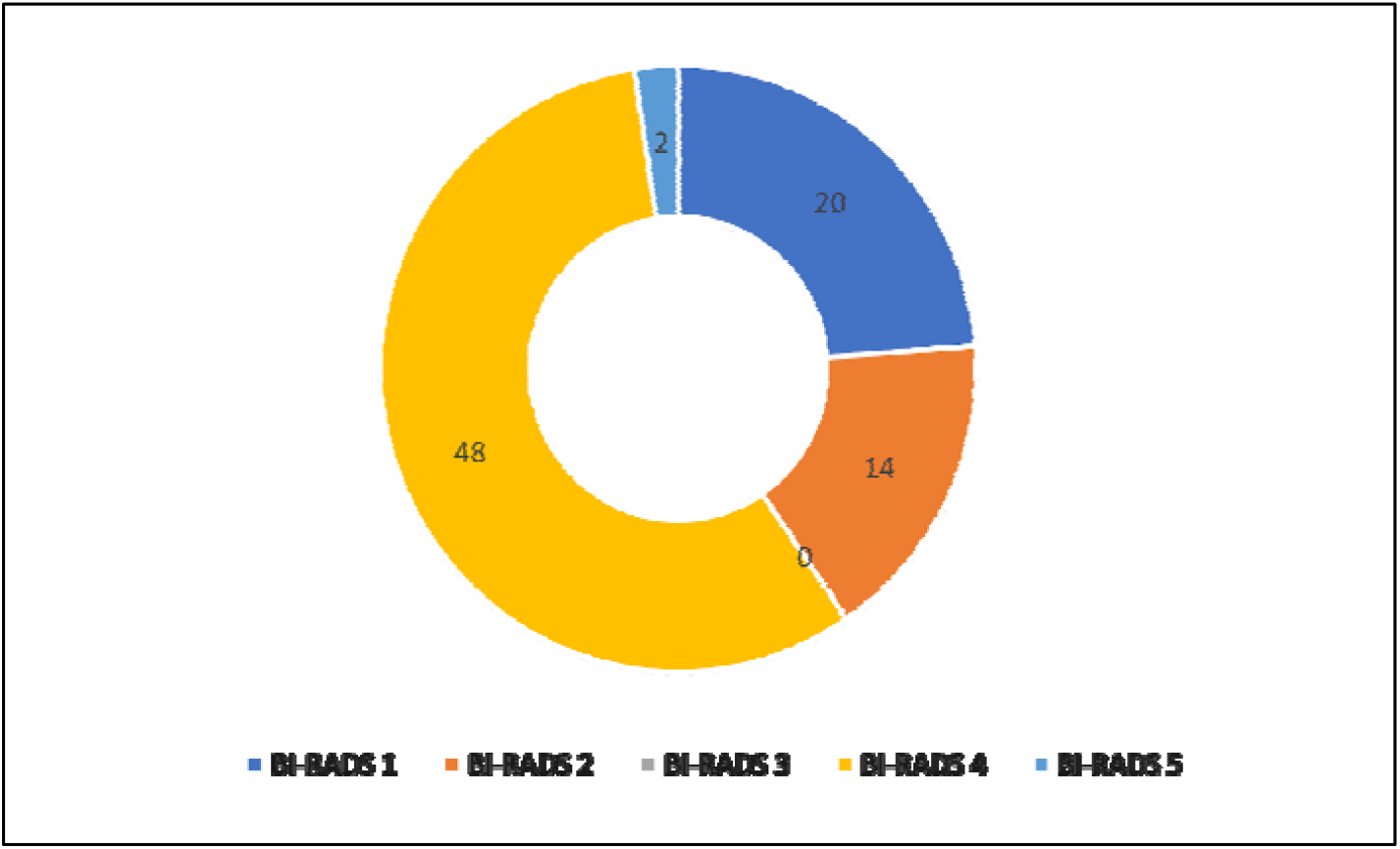
Patient distribution based on BI-RADS.

**Fig. 5:**
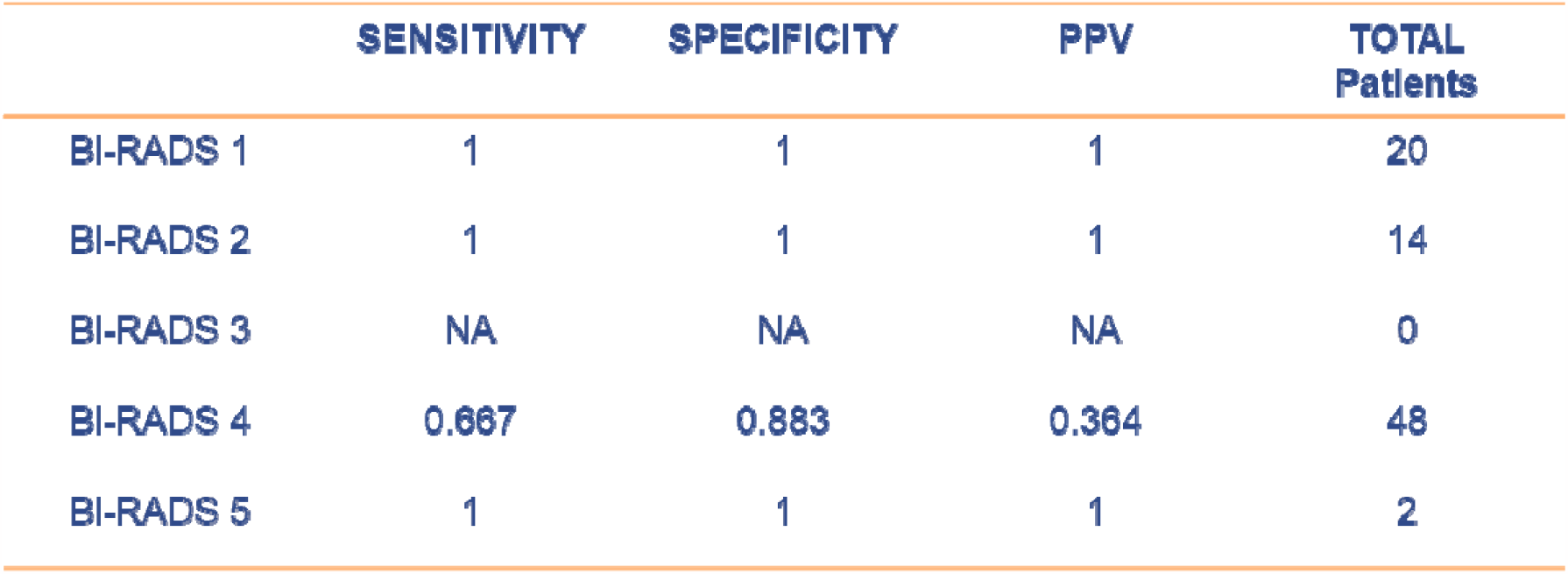
Diagnostic Values of BI-RADS.

**Fig. 6:**
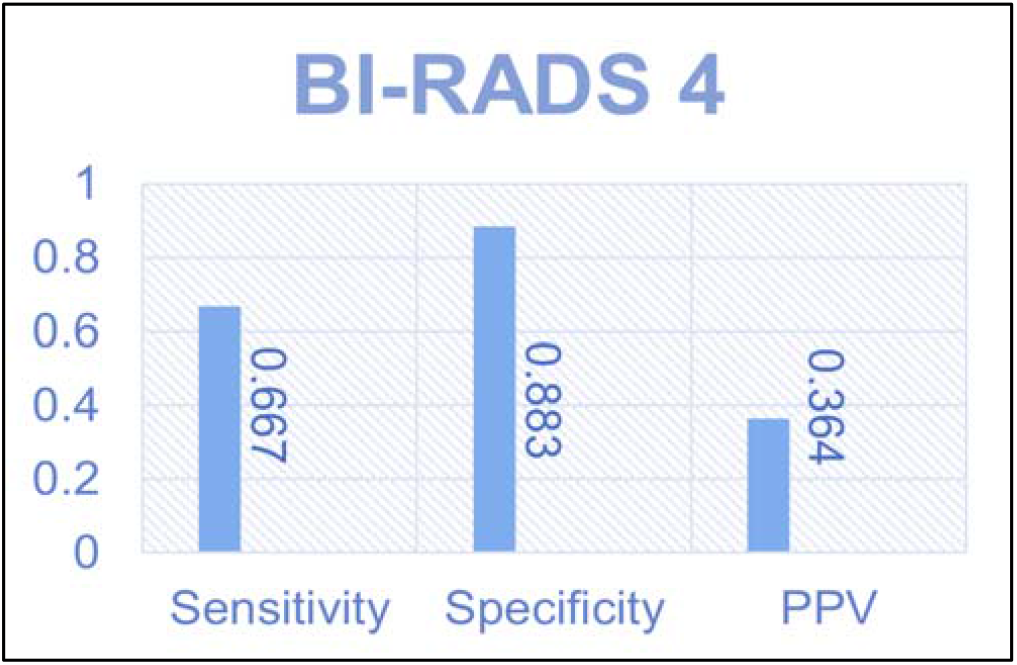
BI-RADS 4 Statistics.

**Fig. 7:**
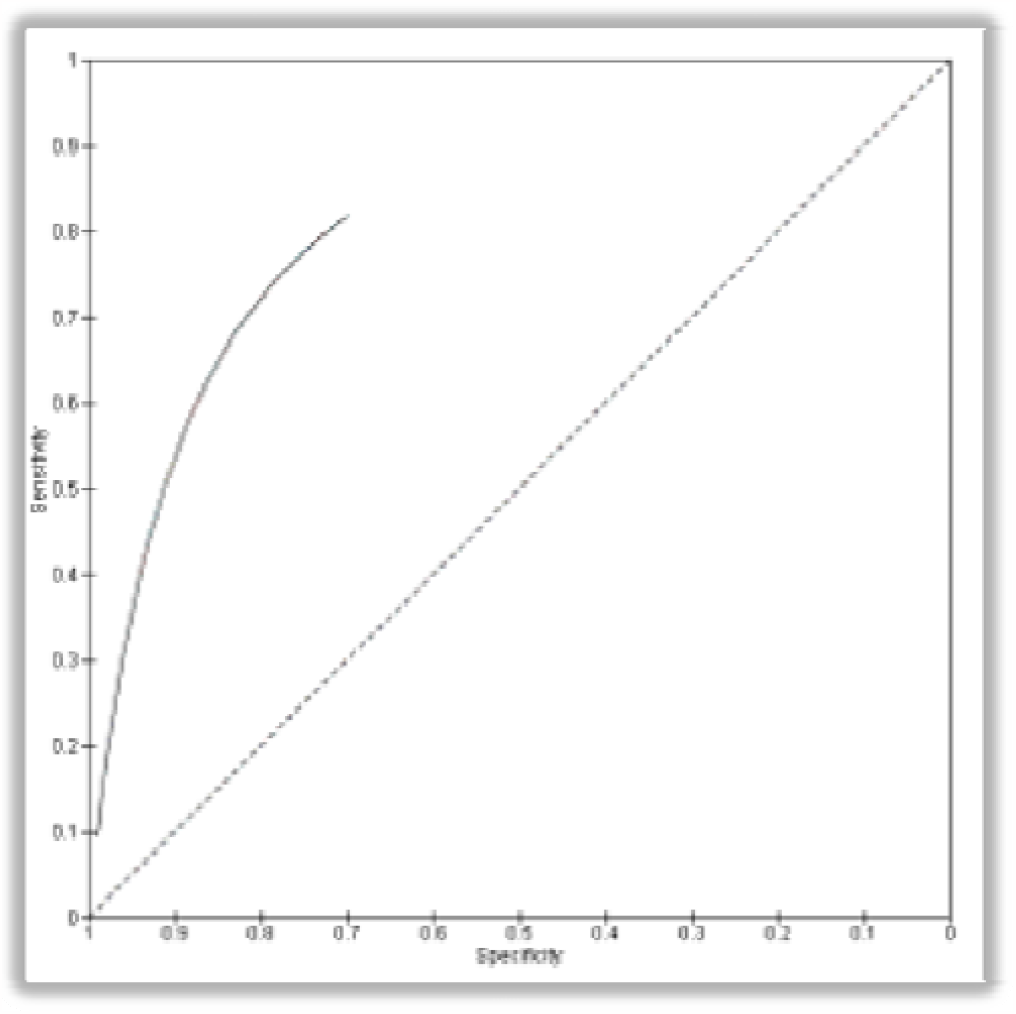
AUC for ROC for BI-RADS 4.

## DISCUSSION

Breast cancer is a potentially life-threatening condition affecting every 1/8th of females worldwide[1], thus its early detection and prompt management are crucial to improving a patient’s condition. For early detection, several radiological modalities are used including Mammography, USG, and MRI in some cases[8]. The findings of this study suggest that USG is a sensitive imaging modality for breast cancer detection. Its high sensitivity makes it a valuable tool, especially in situations where mammography may yield inconclusive results, such as in women with dense breast tissue[4] and in Pregnant females with breast lumps or painful abnormalities. USG can help identify additional lesions that may have been missed by mammography alone.

BI-RADS (Breast Imaging-Reporting and Data System) is a risk assessment and assurance tool developed by the American College of Radiology that provides a widely accepted reporting schema for imaging of the breast. It applies to Ultrasound, Mammography, and MRI.

BIRADS plays a crucial role in standardizing the reporting and interpretation of breast imaging findings. It helps healthcare providers communicate effectively, stratify risk, make appropriate management decisions, establish surveillance protocols, and contribute to quality assurance and research efforts.

**Table 2:** BI-RADS ASSESSMENT CATEGORIES.

| **<u>BI-RADS ASSESSMENT CATEGORIES</u>** |  |
| --- | --- |
| Category 0 | **Incomplete** (Need Additional imaging evaluation) |
| Category 1 | **Negative** |
| Category 2 | **Benign** |
| Category 3 | **Probably Benign** |
| Category 4 | **Suspicious**<br>• Category **4A**: Low Suspicion of Malignancy<br>• Category **4B**: Moderate suspicion of Malignancy<br>• Category **4C**: High Suspicion of Malignancy |
| Category 5 | **Highly Suggestive of Malignancy** |
| Category 6 | **Known Biopsy-Proven Malignancy** |

This study found that Patients whose USG shows Benign growth or can be diagnosed in BI-RADS 1, 2, and 5 can be counted as accurate and precise. It shows that the accuracy of USG BIRADS in classifying the lesion in categories 1, 2, and 5 is high with a sensitivity and specificity of 100%. When the USG diagnosis describes the patient to be in BI-RADS 4, the sensitivity and PPV show poor results due to the inclusion of a wide spectrum of the lesion including inflammatory lesions, breast abscesses, and hyperplasia showing a very low probability of the patient being truly positive when the diagnosis gives a positive result. Hence HPE is recommended for confirmation.

The positive predictive value (PPV) of USG BIRADS 4, which indicates the probability of a positive USG BIRADS IV result being a true positive, was reported at 36.4%. This implies that approximately 36.4%. of the breast lesions identified as suspicious on USG BIRADS 4 are confirmed to be malignant after further evaluation. On the other hand, the negative predictive value (NPV), reflecting the likelihood of a negative USG BIRADS 4 result being a true negative, was estimated at 94%. This indicates that USG has a high ability to rule out breast cancer in cases where no suspicious findings are detected. The diagnostic accuracy of USG BIRADS 4 was 81% in our study. However, it’s worth noting that diagnostic accuracy varies depending on several factors, including the experience of the radiologist, the equipment used, and the characteristics of the patients and lesions being evaluated.

However, the relatively lower specificity of USG BIRADS 4 indicates that it may generate false-positive results, leading to unnecessary further investigations or biopsies. BI-RADS 4 provided more precise categories of breast cancer risk by creating three sub-categories: category 4A includes lesions with low suspicion of malignancy. Category 4B concerns lesions that have an intermediate level of suspicion of malignancy. Follow-up and histological correlation are of great importance in this subgroup because the expected result can be as benign as malignant. Category 4C includes moderately suspicious lesions without, however, classic signs of malignancy. However, this sub-categorization in BIRADS 4 requires clinical knowledge and experience and it recommended that trainee radiologists or radiologists in primary health care centers should emphasize the importance of integrating USG findings with other clinical information and imaging modalities to ensure accurate diagnosis and avoid overtreatment. The results of this study were by the study conducted on a total of 2817 patients which stated PPV FOR BIRADS 5 at 99.6% by Fu C et al.[9] but they did not show the values of sensitivity and specificity. In contrast to this, there were papers by E. Lazarus et al.[10] in which 381 patients were enrolled and A. Guennoun et al. [11] in which 181 patients were enrolled which showed poor quality for BIRADS 3 and 4. We also found a paper that stated a higher diagnostic accuracy of BIRADS 4 than what we found authored by Hye-Jeong Lee et al. [12] with a total of patients 136 in the study.

## Conclusion

This study highlights the diagnostic accuracy of Ultrasonography BI-RADS in detecting breast lesions along with the categorization of lesions as Benign lesions (i.e., BI-RADS 1, 2) and malignant lesions (i.e., BI-RADS 5) with 100% sensitivity and specificity. However for BI-RADS IV, the sensitivity and positive predictive values are relatively low, indicating the need for further confirmation through histopathological examination as BI-RADS 4 includes a wide spectrum of breast lesions. This study also shows the importance of clinical aspects along with imaging methods to improve accuracy, especially with a wide spectrum of breast lesions. The pathological analysis of the radiological aspect allows a good diagnostic approach; however, the Histo-pathological examination remains the gold standard examination to determine the benign or malignant nature of the breast tumor. These study results help in understanding the role of USG in breast cancer diagnosis and the significance of BI-RADS in standardizing imaging reports.

## Data Availability

All data produced in the present study are available upon reasonable request to the authors

## Author contributions

All authors have read and approved the final version of the manuscript.

